# BUDGET IMPACT ANALYSIS OF THE INTRODUCTION OF RANIBIZUMAB’S BIOSIMILAR TO THE JORDANIAN JOINT PROCUREMENT SYSTEM

**DOI:** 10.64898/2026.01.13.26344067

**Authors:** Jalal Abu Hamida, Nimer S. Alkhatib, Khawla Abu-Hammour, Shiraz Halloush, Amanj Baker, Bander Balkhi, Osama Alfayez, Rimal Mousa

## Abstract

**Introduction:** The adoption of biologic therapies imposes a substantial financial burden on the Jordanian healthcare system. Ranibizumab is prescribed for various retinal disorders, and its associated costs are considerable. The introduction of biosimilars is beneficial in retaining desired clinical parameters while providing cost relief and enhanced access to patients.

**Objective:** To examine the budget impact and expanded access of switching to ranibizumab’s biosimilar for the management of retinal diseases in guideline-based practice and in real-world practice in Jordan.

**Method:** A 4-year budget impact analysis from Jordanian public sector payer’s sector was performed (2023 to 2026) that included patient prevalence and incidence, average ranibizumab dose per year, and anticipated shifts in the market share of ranibizumab and aflibercept. The model took into account the anticipated price erosion of the biosimilar in 2025 and 2026. Sensitivity analyses were performed to assess the effect of changes in uptake rates, price, and market share.

**Results:** The annual cost savings per patient when switching from aflibercept to ranibizumab’s biosimilar were from 20.55 JOD (Jordanian Dinar) to 1519.93 JOD, translating to a percentage saving of 2.68% to 35.12% across the various scenarios and indications. The total budget impact ranged widely from 6.9 M JOD to 21.2 M JOD based on treatment regimens adjusted to current practice, PRN (Pro re nata), or T&E (Treat and extend). Patient access improved between 2.75% to 124.76% in the different scenarios.

**Conclusion:** The introduction of ranibizumab’s biosimilar significantly reduces the expenditures and enhances treatment access.

## Introduction

Age-related macular degeneration (AMD), diabetic macular edema (DME), and retinal vein occlusion (RVO) are the leading causes of vision-threatening conditions globally (Tsiara 2020). Such diseases represent a tremendous burden on patients and on their healthcare systems, especially in resource-limited middle-income countries, such as Jordan, where budget constraints and resource shortages are part of daily practice (Hariprasad et al., 2025). In Jordan, there are challenges associated with retinal disorders, as most of the patients experience difficulties in seeking early treatment. The availability of specialized ophthalmologic care in the rural areas, shortage of healthcare providers (HCPs), and early detection programs which impact the early diagnosis and treatment of retinal disorders ((IAPB), 2025; Al-Dwairi et al., 2024). Delays in diagnosis because of access problems can negatively impact treatment outcomes. Wider outreach and affordable treatments, like ranibizumab’s biosimilars, may help more patients get timely care to save their sight (Al-Dwairi et al., 2024). The number of patients with AMD, DME, and RVO is escalating globally (Al-Dwairi et al., 2024; Klein et al., 1984; Rehak & Wiedemann, 2010; Song et al., 2019; Wong et al., 2014), Roughly 10-15% of AMD cases are categorized as the “wet” or exudative type, also referred to as neovascular macular degeneration (nAMD) (Gehrs et al., 2006). The treatment of retinal diseases is costly because it incurs the cost of different treatments modalities, including the cost of anti-VEGF (Vascular Endothelial Growth Factor) injections, laser treatment, imaging, and surgery (Simmonds et al., 2024). Thus, because the demand for these treatments keeps increasing in Jordan, it is essential to explore cost-saving strategies that improve access without sacrificing effectiveness. the biosimilars available in the form of ranibizumab could present an attractive, cost-effective treatment choice in the anti-VEGF family in the field of ophthalmology (Cano et al., 2025). However, before these can be rolled out nationally, we need to thoroughly evaluate their economic impact within the Jordanian healthcare system. Budget impact analysis (BIA) plays an important role in the evaluation process aimed at measuring the short- and medium-term cost implications of the treatment regimen within the healthcare settings (Sullivan et al., 2014). BIA considers affordability, budget sustainability, and resource planning, all vital in the delivery and operation of health systems such as Jordan, where finances are limited and there has been an increase in the need for expensive biologics (Almaaytah, 2020). The purpose of this proposed research work is to examine the budget impact and increased accessibility with the use of the ranibizumab’s biosimilar in the treatment of nAMD, DME, and RVO in the Jordanian healthcare system. The proposed model will outline the financial projections in using the biosimilar in the Jordanian healthcare system over a four-year duration and hence make valuable recommendations.

## Methodology

### Model Framework

A static, economic BIA was conducted using Microsoft Excel, following the International Society for Pharmacoeconomics and Outcomes Research (ISPOR). The analysis adopted the public payer perspective of the Jordanian General Procurement Department (GPD), and only direct drug acquisition costs were included in the analysis. No other cost components were considered. The time horizon selected is four-years (2023-2026), which encompasses the usual duration of care required to treat chronic retinal conditions (3 to 5 years) and also enables policymakers to project budget impact over multiple years, which helps to estimate the budget impact on finances (Faleiros et al., 2022; Heier, Korobelnik, et al., 2016).

As patient-level data was unavailable, the model relied on aggregated national consumption data and expert consensus for utilization rates. Treatment mix was defined from hospital records, HCPs, pharmacies, and GPD reports. The model represented four key scenarios across all of their indication, guideline-based utilization which assumes use of pro re nata (PRN) and treat and extend (T&E) treatment regimen based on international scientific literature and current practice utilization which represents the actual pattern use of in clinical practice in Jordan, each scenario was modeled in both with and without the availability of ranibizumab’s biosimilar as shown in **(Figure 1)**. The costs and quantities for aflibercept, reference ranibizumab, and its biosimilar were acquired as inputs from GPD, as shown in Table 1. The GPD undertakes centralized purchase of drugs for the entire public health system in Jordan. It was mainly serving the Ministry of Health, the Royal Medical Services, and university hospitals (Statistics, 2017).

**Table 1.** Anti-VEGFs consumption quantity and cost per vial between 2021 to 2024 based on GPO data.

| **Table 1 \| Anti-VEGFs consumption quantity and cost per vial between 2021 to 2024 based on GPD data.** |  |  |  |  |  |  |
| --- | --- | --- | --- | --- | --- | --- |
| Year | Aflibercept quantity (Vial) | Aflibercept Cost per vial (JOD) | Reference ranibizumab quantity (Vial) | Reference ranibizumab cost per vial (JOD) | Ranibizumab's biosimilar quantity (Vial) | Ranibizumab's biosimilar cost per vial (JOD) |
| 2021 | 4,450 | 559.01 | 3,060 | 540.18 | - | - |
| 2022 | 3000 | 559.01 | 2320 | 540.18 | - | - |
| 2023 | 3444 | 508.14 | 3,020 | 497.64 | - | - |
| 2024 | 6125 | 488.60 | 2025 | 317 | 2025 | 317 |

**Abbreviations:** GPD: Governmental Procurement Department, JOD: Jordanian Dinar.

**Figure 1.**
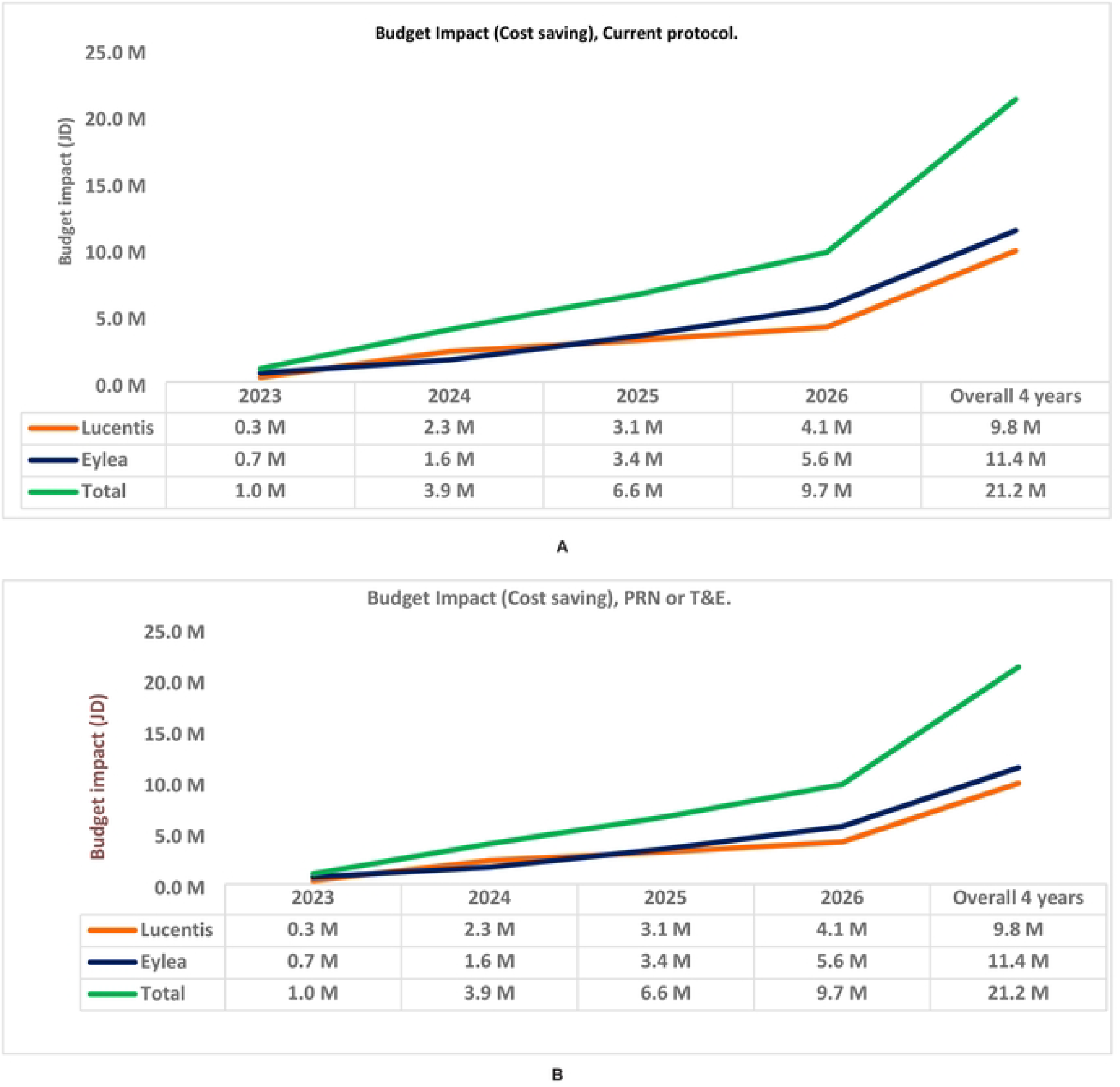
A: Budget impact of ranibizumab’s biosimilar admission in PRN and T&E over 2023 10 2026. B: Budget impact of ranibizumab’s biosimilar admission in Jordanian current practice over 2023 to 2026. **Abbreviations:** M: Million. PRN: Re Nata. T&E: Treat and Extend.

### Population

The model used average treatment costs per hypothetical patient for each indication. The numbers of treated nAMD, DME, and RVO patients were based on national consumption data (2021–2024) from the GPD and was subsequently adjusted by utilization rates, all data were accessed for research purposes from 1/April/2024 to 15/March/2025 and all data were fully anonymized. Ranibizumab (reference and biosimilar) and aflibercept utilization rates were obtained by dividing total annual consumption by the average quantity of vials used per patient per year. The current utilization estimates were informed by expert interviews with HCPs in different institutions. The current usage rate of utilization was two vials per patient per year for patients receiving aflibercept and three vials per patient per year for patients receiving ranibizumab. The patient with DME or RVO in guideline-based scenarios, including PRN or T&E regimens, consumed an average of 7 vials of aflibercept or ranibizumab annually, while the nAMD patient consumed 10 vials of either drug per year (Akbas et al., 2023; Campochiaro et al., 2010; Heier et al., 2012; Volkmann et al., 2020).

### Model Assumptions

The same assumptions were used for all scenarios, including 1) Full adherence by patients in each of the treatment arms. 2) equivalence in efficacy and safety between the biosimilar and the reference treatment based on published clinical studies. 3) the healthcare system has sufficient capacity to meet the projected need with no shortages of facilities or staff.

### Analysis

A comprehensive strategy was implemented to undertake several economic evaluations on the budget impact of ranibizumab’s biosimilar availability. The purpose of these assessments was to estimate the budgetary impact, patient access variations, and relative costs of alternative anti-VEGF treatments in Jordan. The total annual budget required to treat a hypothetical patient was estimated by multiplying the number of vials needed for each product for each indication by the cost per vial. Annual treatment costs for patients with specific eye conditions were compared using a direct subtraction approach. Switching to ranibizumab’s biosimilar was assumed to expand access to anti-VEGF treatment for patients. The increase in access was estimated by dividing the savings generated from switching from a higher-cost drug to a lower-cost alternative by the treatment cost of the higher-cost drug (Alkhatib et al., 2024).

Number needed to convert (NNC) describes the number of patients that need to switch from the high-cost treatment to the lower-cost option to treat 10 patients. NNC is calculated by multiplying the cost of the lower-cost drug by 10 and then dividing the result by the difference in cost between the higher-cost drug and the lower-cost drug (McBride et al., 2020).

To assess the value-for-money of each anti-VEGF treatment, the relative expenditure per JOD (Jordanian dinar) was calculated by comparing the cost of each treatment with the least expensive option.

Deterministic sensitivity analysis was performed to analyze the robustness of the base-case results to changes in parameter values (drug prices, epidemiological values like prevalence and incidence rates, market share, and utilization rates). Data was presented in a tornado diagram, showing the impact of each parameter on the budget impact model.

## Results

### Patient Distribution for Anti-VEGF Therapy by Disease Condition (2021–2026)

The total number of patients treated with anti-VEGF therapy between 2021 and 2024 in Jordan showed a continuous increase. Approximately 70% of the treated population per year were treated for DME. nAMD and RVO accounted for approximately 20% and 10% of cases, respectively. We extrapolated the total patient population for the next two years using the average annual growth rate of 17.26% from 2021 to 2024. The total number of patients increased from 3,245 in 2021 to 6,068 in 2026, with intermediate values of 2,273 in 2022, 2,729 in 2023, 4,413 in 2024, and 5174 in 2025. The market share of vials of aflibercept remained steady around 60% from 2021 to 2026. The remaining 40% belonged to reference ranibizumab before the launch of its biosimilar; after this, the share was divided between the reference product and the biosimilar.

### Treatment Costs and Patient Access

The direct annual budget for the two treatment options was calculated based on two main scenarios, with and without access to the ranibizumab biosimilar. For the biosimilar scenario, we included an average price erosion of 11.4% to represent anticipated decreases in drug prices over time. In a no-biosimilar scenario, however, roughly 0% price erosion was modelled (stable drug price without market competition). The cumulative treatment cost without the biosimilar in the current practice scenario from 2023 to 2026 is 23.47 million (M). JOD, while in the case of the existence of the biosimilar, the cumulative cost was 16.59 M JOD. The cumulative treatment cost from 2023 to 2026 in the PRN or T&E scenario without the biosimilar is 77.27 M JOD, while in the case of the existence of the biosimilar, the total cumulative cost was 56.05 M JOD. The introduction of ranibizumab’s biosimilar in the current practice could decrease the annual cost for each patient who takes aflibercept from 102 JOD to 351 JOD and for each patient who takes ranibizumab from 128 JOD to 874 JOD, while in PRN or T&E, the annual saving for each patient who takes aflibercept is from 356 JOD to 1757 JOD and for each patient who takes ranibizumab from 297 JOD to 2915 JOD, based on the indication and year. Switching patients who take aflibercept to ranibizumab’s biosimilar could decrease the annual cost of treatment in the current practice from 20.55 JOD to 23.20 JOD, while in the PRN or T&E scenario, it decreases the annual cost from 942.41 JOD to 1,519.93 JOD.

In addition to the direct cost reductions, switching patients to ranibizumab’s biosimilar was estimated to improve patient access to the treatment from 2.75% to 117.19% in the current practice, and from 54.13% to 124.76% in the PRN or T&E scenarios. This is indicative of giving access to more patients with the same budget when transitioning from aflibercept and reference ranibizumab.

### Number Needed to Convert (NNC)

In any case examined, ranibizumab’s biosimilar was the most affordable alternative compared with aflibercept and reference ranibizumab. To treat another 10 patients with ranibizumab within the same budget, about 18–19 patients would have to be switched from aflibercept to the biosimilar in the PRN or T&E scenarios in either 2025 or 2026. In the current practice scenario, about 363 patients would have to be switched to have the same gain of patient access.

### Relative Value of the Jordanian Dinar (JOD)

Relative expenditure of ranibizumab’s biosimilar versus aflibercept and reference ranibizumab was between 1.03 and 2.17 in the current practice scenario, and between 1.54 and 2.25 in the PRN or T&E scenario. This reflects the reduction in overall treatment cost when other anti-VEGFs are switched to the biosimilar, highlighting its role in lowering overall treatment costs in the different treatment scenarios.

### Total Budget Impact of Ranibizumab’s Biosimilar

From 2023 to 2026, the entry of ranibizumab’s biosimilar affected the prices of eye treatment drugs in Jordan. Price reductions started after registration of the biosimilar by the JFDA in 2022, which led to a 7.88% reduction in list price for ranibizumab and 9.10% for aflibercept, even before its inclusion in tenders. Following the addition to the official tender, price erosion reached 36.30% for the biosimilar and 3.85% for aflibercept, yielding substantial cost savings prior to any exclusion. In 2023, the total treatment cost was 8.54% lower, which accounted for 303,653 JOD. In 2024, savings increased to 23.79%, accounting for 1.3 M JOD. Between 2025 and 2026, the reductions are expected to be as high as 32.5% and 40.21%, accounting for 2.1 M JOD and 3.1 M JOD, respectively.

The total budget impact of the biosimilar adoption over 4 years is projected to range between 6.9 M JOD in the current practice and 21.2 M JOD in PRN or T&E protocols, as shown in (**Figure 2)**, demonstrating a significant reduction in costs in conjunction with the biosimilar’s entry to the market.

**Figure 2.**
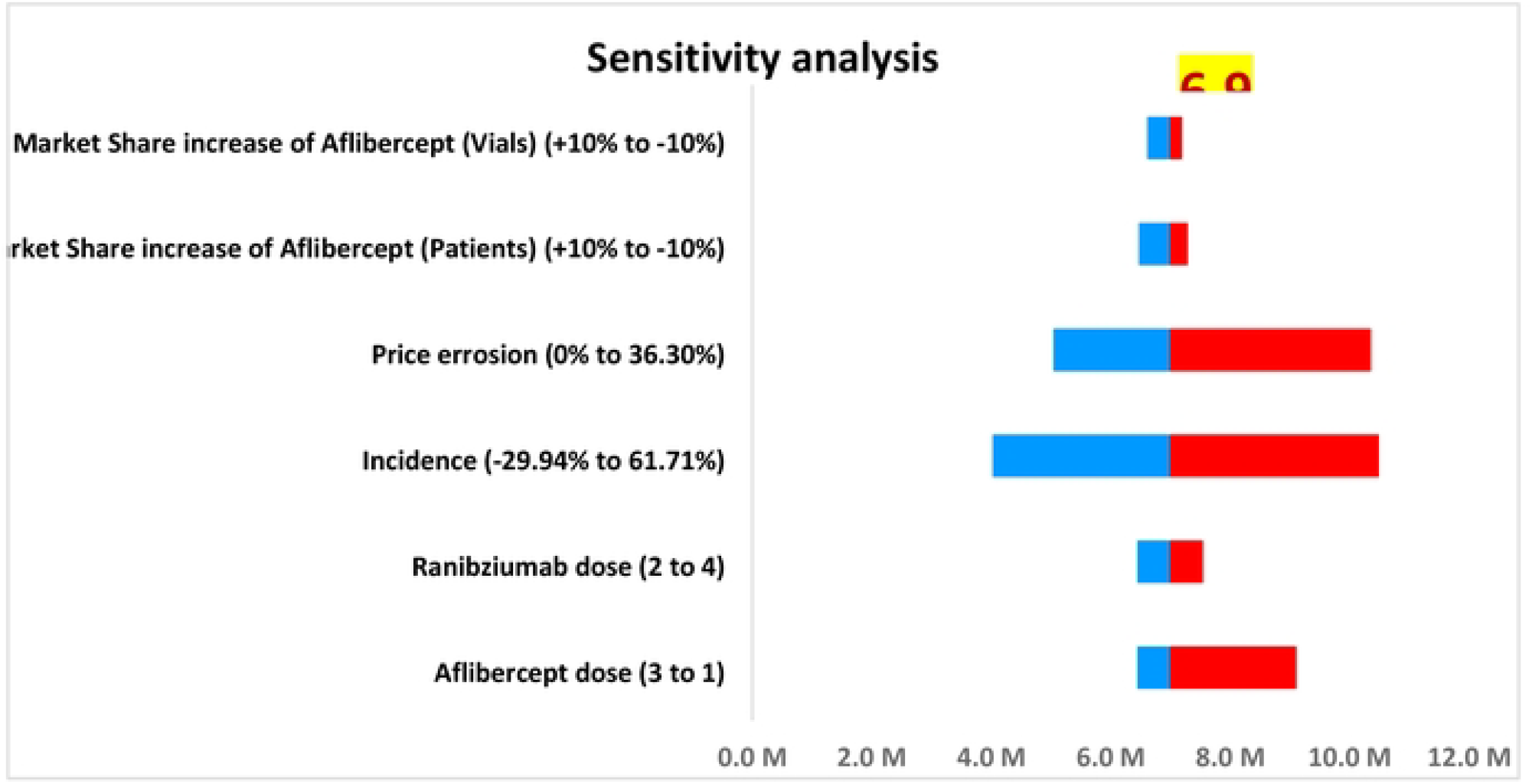
Tornado chart representing the deterministic sensitivity analysis of key parameters influencing the budget impact of introducing ranibizumab’s biosimilar under Jordanian current practice. **Abbreviations**: M: Million.

### Sensitivity analysis

The sensitivity analysis as demonstrated in (**Figure 3**) indicated that the 4-year budget impact for ranibizumab biosimilars was sensitive to variation in the key input parameters. Changes in the dose of aflibercept had a significant impact on the BIA, varying from 6.45 M JOD to 9.11 M JOD, compared to the base case of 6.9 M JOD. Ranibizumab dose changes led to an intermediate fluctuation, ranging from 6.45 M JOD to 7.55 M JOD. Differential rate of treated eye conditions led to a larger spread of the budget, with cost estimates ranging from 4 M JOD to 10.49 M JOD. Price erosion cases also showed large variation, with the budget being from 5 M JOD when there is no price erosion to 10.36 M JOD when the price is eroded to maximum. Shifts in market share for aflibercept led to a budget variability of 6.47 M JOD to 7.29 M JOD depending on the number of patients. Likewise, the revenues for different shares from the market according to the vial used were between 6.61 M JOD and 7.19 M JOD. These results are visually represented in the tornado diagram, which shows the relative impact of each variable on the total budget estimation and ensures that findings were robust under all tested scenarios and assumptions.

## Discussion

Ocular diseases such as AMD, DME, and RVO have a significant burden on health systems. In daily clinical practice, Jordan faces significant constraints and resource shortages, making the economic burden of treating these retinal diseases a significant concern. The current investigation aimed to assess the budget impact and expanded access of switching to ranibizumab’s biosimilar for the management of retinal diseases in both guideline-based and real-world practice in Jordan. The four-year BIA showed that switching to the biosimilar could save between 6.9 and 21.2 million JOD compared with continuing other therapies. The cost reduction observed can help the health system manage the growing number of patients with retinal diseases without increasing the total budget. Moreover, adopting the biosimilar would allow more patients to receive therapy under the same financial resources. The results of this BIA suggest that the introduction of ranibizumab’s biosimilar is a practical and affordable approach.

The findings of this BIA are consistent with international and regional evidence (Brown et al., 2020; Carrasco et al., 2020; Dakin et al., 2014; El-Dahiyat & Eljilany, 2020; Hernandez et al., 2018; Holekamp et al., 2020; Izidoro et al., Impacto orçamentário da incorporação de medicamentos para tratamento em segunda linha do edema macular diabético no SUS sob a perspectiva da Secretaria de Estado de Saúde de Minas Gerais, Brasil./2019; Ross et al., 2016; Samacá-Samacá et al., 2025; Stein et al., 2014). Reports have shown that the cost of anti-VEGF therapy can reduce treatment expenditure without compromising clinical outcomes. For instance, in the United Kingdom, Dakin and colleagues demonstrated that ranibizumab and bevacizumab achieved comparable improvements in visual acuity, with lower-priced bevacizumab leading to marked savings (Dakin et al., 2014). The results of this study highlight the economic importance of using equally effective but less costly options. A study conducted in the United States by Stein et al. reported that ranibizumab was not cost-effective compared with bevacizumab for nAMD (Stein et al., 2014). On the other hand, Heier et al. confirmed that all three major anti-VEGF agents, namely aflibercept, ranibizumab, and bevacizumab, provided similar visual benefits in patients with DME (Heier, Bressler, et al., 2016). This information reinforces that price rather than efficacy largely determines the economic burden of anti-VEGF therapy.

Real-world evaluations from other markets have also confirmed that economic efficiencies can be achieved without sacrificing outcomes. A study in France found both aflibercept and ranibizumab to be cost-effective in routine practice. These results were largely influenced by dosing frequency and acquisition cost (Carrasco et al., 2020). Similarly, Samacá-Samacá et al. showed that market competition and introduction of biosimilars such as faricimab resulted in major national budget savings while maintaining efficacy (Samacá-Samacá et al., 2025). These studies support that introducing a ranibizumab biosimilar in Jordan follows a global movement toward affordable retinal treatment. This effort ensures positive patient outcomes and reduces overall healthcare spending.

From a regional perspective, adopting Ranibizumab’s biosimilar in Jordan is supported by similar findings from other Middle Eastern and middle-income healthcare systems. A recent Saudi Arabian study by El-Dahiyat and Eljilany showed that substituting aflibercept for ranibizumab could achieve 9% to 51% cost savings across retinal indications (El-Dahiyat & Eljilany, 2020). This result surely demonstrates how pricing competition can significantly reduce expenditure (El-Dahiyat & Eljilany, 2020). Moreover, a Brazilian study estimated that public sector adoption of lower-cost anti-VEGF agents could reduce total expenditures for DME management by nearly one-half (Izidoro et al., Impacto orçamentário da incorporação de medicamentos para tratamento em segunda linha do edema macular diabético no SUS sob a perspectiva da Secretaria de Estado de Saúde de Minas Gerais, Brasil./2019).

The consistent findings across international and regional studies suggest that incorporating ranibizumab’s biosimilar into national procurement systems represents a timely and evidence-based policy decision. For Jordan, the anticipated budget relief could be reinvested to improve screening, increase treatment, and improve overall ophthalmic services in public hospitals. Comparable results from Colombia and France further suggest that the availability of biosimilars and newer agents creates a competitive environment that ultimately benefits health systems through lower cost and increasing treatment opportunities for more patients (Carrasco et al., 2020; Samacá-Samacá et al., 2025). The successful implementation of biosimilar therapy, however, will depend on coordinated regulatory oversight, clinician education, and transparent procurement processes. Validating and improving these findings in the Jordanian context will require ongoing observation of clinical outcomes, patient satisfaction, and real-world utilization after biosimilar adoption.

This analysis has some limitations that should be considered. First, the model relied on secondary data rather than patient-level cost or utilization data. Variations in clinical practice patterns and injection frequency may influence actual expenditures. The analysis assumed therapeutic equivalence of treatments based on evidence from large comparative trials. Differences in adherence, dosing intervals, and physician preferences may affect treatment outcomes. The model did not include indirect costs such as productivity loss or caregiver burden. Finally, the projections represent short-to-medium-term estimates. Continuous monitoring of real-world utilization, clinical outcomes, and patient satisfaction following biosimilar adoption will be important to validate and refine these findings in the Jordanian context.

## Conclusion

The introduction of ranibizumab’s biosimilar represents one of the most important steps taken by Jordanian authorities in managing retinal diseases. This biosimilar has contributed not only to the emergence of an inexpensive alternative for patients but also to saving budgets on the healthcare system by influencing the current prices of anti-VEGF therapies.

## Statements and Declarations

### Authors Contributions

Jalal Abu Hamida, Rimal Mousa, Nimer S Akhatib carried out the study conception, design, data collection, analysis, budget impact modeling, and manuscript drafting. Bander Balkhi. and Khawla Abu-Hammour, Amanj Baker, Shiraz Hallous. contributed exclusively to the critical review and revision of the manuscript. All authors read and approved the final manuscript

### Data availability

Data will be available upon request

### Compliance with Ethical Standards

Ethical approval to obtain the information needed to conduct this study was obtained from the Institutional Review Board of the MOH research committee. No information was collected directly from patients and thus, consent forms were not collected on the patient level.

## Acknowledgement

Ongoing Research Funding Program (ORF-2025-76), King Saud University, Riyadh, Saudi Arabia

